# Development of a comprehensive cardiovascular disease genetic risk assessment test

**DOI:** 10.1101/2024.05.06.24306379

**Authors:** Laura M. Amendola, Alison J. Coffey, Josh Lowry, James Avecilla, Alka Malhotra, Aditi Chawla, Stetson Thacker, Julie P. Taylor, Revathi Rajkumar, Carolyn M. Brown, Katie Golden-Grant, Rueben Hejja, Jennifer A. Lee, Phillip Medrano, Becky Milewski, Felipe Mullen, Andrew Walker, Adriana Huertez-Vasquez, Mauro Longoni, Denise L. Perry, Damon Hostin, Subramanian S. Ajay, Akanchha Kesari, Samuel P. Strom, Elliott Margulies, John Belmont, David E. Lanfear, Ryan J. Taft

## Abstract

**Background:** Despite monogenic and polygenic contributions to cardiovascular disease (CVD), genetic testing is not widely adopted, and current tests are limited by the breadth of surveyed conditions and interpretation burden.

**Methods:** We developed a comprehensive clinical genome CVD test with semi-automated interpretation. Monogenic conditions and risk alleles were selected based on the strength of disease association and evidence for increased disease risk, respectively. Non-CVD secondary findings genes, pharmacogenomic (PGx) variants and CVD polygenic risk scores (PRS) were assessed for inclusion. Test performance was modeled using 2,594 genomes from the 1000 Genomes Project, and further investigated in 20 previously tested individuals.

**Results:** The CVD genome test is composed of a panel of 215 CVD gene-disease pairs, 35 non-CVD secondary findings genes, 4 risk alleles or genotypes, 10 PGx genes and a PRS for coronary artery disease. Modeling of test performance using samples from the 1000 Genomes Project revealed ∼6% of individuals with a monogenic finding in a CVD-associated gene, 6% with a risk allele finding, ∼1% with a non-CVD secondary finding, and 93% with CVD-associated PGx variants. Assessment of blinded clinical samples showed complete concordance with prior testing. An average of 4 variants were reviewed per case, with interpretation and reporting time ranging from 9-96 min.

**Conclusions:** A genome sequencing based CVD genetic risk assessment can provide comprehensive genetic disease and genetic risk information to patients with CVD. The semi-automated and limited interpretation burden suggest that this testing approach could be scaled to support population-level initiatives.

## Introduction

Identifying the genetic underpinnings of cardiovascular disease (CVD), including both monogenic and polygenic factors, can improve patient risk stratification, refine clinical diagnoses, and inform targeted treatment opportunities^1–4^. Guidelines recommend genetic testing for patients presenting with cardiomyopathy, arrhythmias, dyslipidemias, and aortopathies, but such testing is substantially underutilized. For example, a recent retrospective cohort study found that only 0.8% to 1.6% of patients newly diagnosed with CVD, and for whom guidelines recommended testing, received any kind of genetic investigation^5^. For those patients who do receive genetic testing, the results may be affected by inter-laboratory variability in assay differences (e.g. panel vs exome), the genes selected for interrogation, variant interpretation and variant classification^6^. Inconsistent testing practices in the CVD population can exaggerate inequities in access to a genetic diagnosis, undermine the value of genetic testing, and reduce the likelihood of reimbursement.

A growing body of evidence supports the use of genome sequencing (GS) as a first-line test, particularly in the neonatal acute care population^7^ and in children with signs and symptoms of a genetic disease ^8^. We reasoned that a GS-based CVD-focused genetic risk assessment test may simplify CVD genetic testing practices by providing a single, multifaceted, high-throughput testing platform for patients with a CVD phenotype for which genetic testing is recommended. Here we describe the development of the TruGenome^TM^ CVD Test including monogenic CVD and risk allele evaluations, the selection of evidence-supported pharmacogenetic (PGx) alleles, identification of a population-sensitive polygenic risk score (PRS), and approaches to reduce test interpretation and reporting time. We also report the frequency of findings in unrelated individuals in the 1000 Genomes Project cohort, providing an estimate of CVD risk and enabling an assessment of test outcomes across multiple genetic ancestries. Finally, we explore test interpretation burden and compared findings from the TruGenome^TM^ CVD Test to findings in a set of clinical samples from 20 previously tested individuals with CVD. The approach and outcomes reported here may inform future integration of GS applications in the diagnosis and management of CVD and other disease areas.

## Methods

Full methods are available in the Supplemental Materials. The data that support the findings of this study are available from the corresponding author upon request. Institutional review board approval for activities involving human samples was received from WIRB-Copernicus Group (Protocol 20241866). Informed consent was not required.

## Results

### Design of the TruGenome^TM^ CVD Test

Test development focused on CVD phenotypes with a high prior probability of a genetic etiology and relevance in an adult cardiac clinic population, particularly aortopathy, arrhythmia, cardiomyopathy, dyslipidemia, coronary artery disease (CAD), hypertension and thrombophilia. The final test design includes five components: a CVD gene panel, select CVD risk alleles, non-CVD secondary findings, a PRS for CAD, and PGx findings. Each component integrates reporting thresholds set to support the return of high confidence, medically actionable genetic information (**Figure 1**).

**Figure 1.**
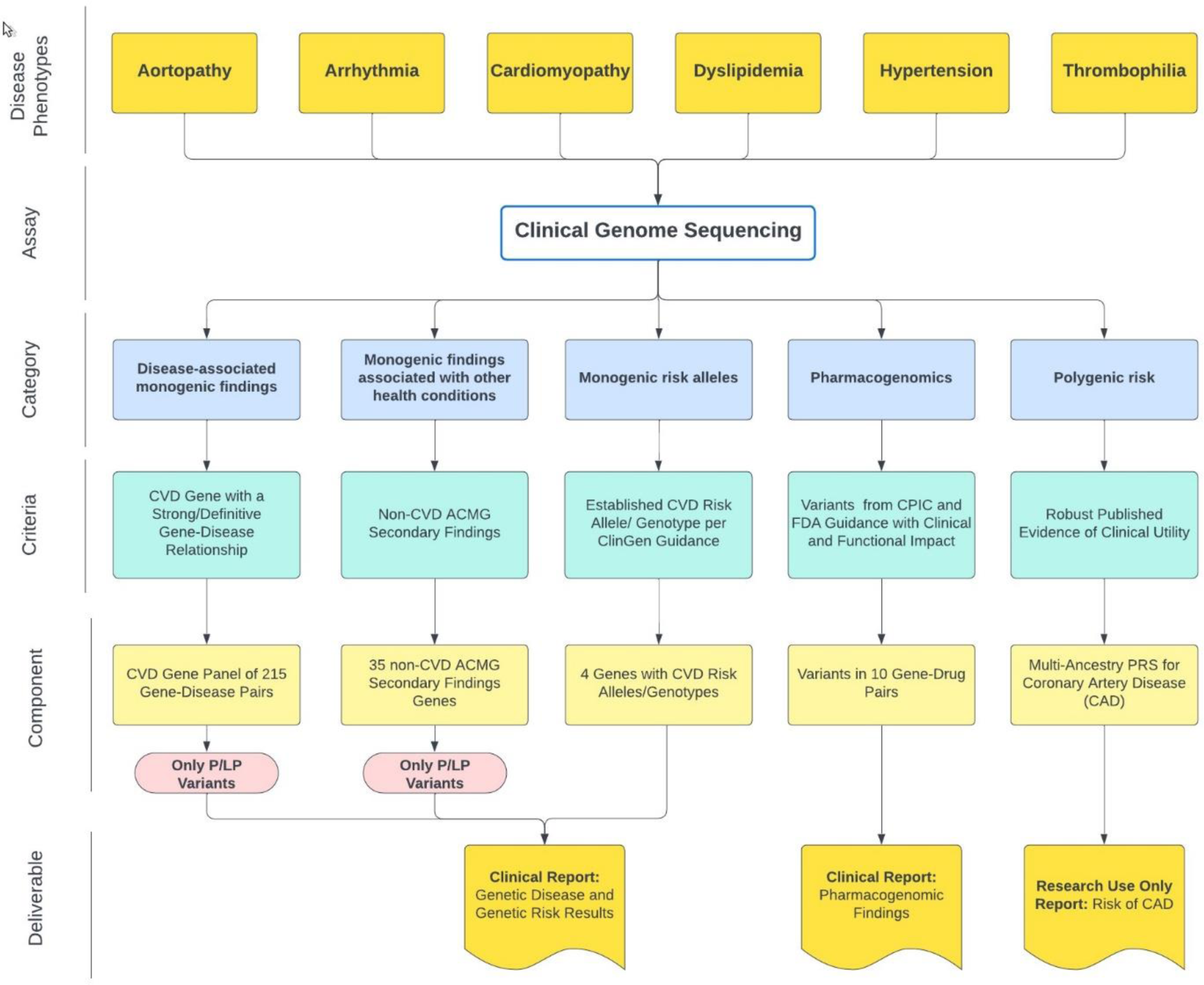
TruGenome^TM^ CVD test design and reportables. Test design focused on CVD phenotypes relevant to cardiac clinic patients. Monogenic conditions and risk alleles were evaluated for the strength of evidence for increased disease risk. CVD PRSs and PGx findings were also explored for inclusion. The test includes five components: a 215 gene-disease pair CVD panel, the 35 remaining non-CVD ACMG secondary findings genes, 4 risk alleles or genotypes, PGx findings across 10 gene-drug pairs and a PRS for CAD. Test deliverables include a clinical report for genetic disease and genetic risk findings, an RUO CAD PRS report and a clinical report of pharmacogenomic findings.

#### Cardiovascular Disease Gene Panel

Three hundred and thirty-nine gene-disease pairs related to the relevant CVD phenotypes were identified from the literature, commercial gene panels, and publicly available gene-disease relationship (GDR) databases. One hundred and thirty nine of these 339 gene-disease pairs were included without further assessment based on a definitive or strong (D/S) GDR classification from ClinGen, Illumina Clinical Services Laboratory (ICSL) and groups that use the ClinGen Gene-Disease Validity framework. Forty-five gene-disease pairs were excluded based on a refuted or disputed GDR from ClinGen, ICSL or a group that uses the ClinGen framework. Using an expedited curation methodology (see Supplemental Methods) the remaining 155 gene-disease pairs were assessed and an additional 76 gene-disease pairs were included, yielding a final panel of 215 gene-disease pairs (**Supplemental Figure 2, Supplemental Table 2**).

Cardiomyopathy was the most common CVD phenotype amongst the gene-disease pairs (66/215, 30%). Fifty GDRs (23%) were associated with more than one CVD phenotype, the majority of which included cardiomyopathy and arrhythmia in the phenotypic spectrum of disease (36/50). Curation of gene-disease pairs without a publicly available or ICSL GDR classification led to the inclusion of 16 gene-disease pairs, associated with cardiomyopathy (7 genes), dyslipidemia (4 genes), hypertension (4 genes), and arrhythmia (1 genes). Additionally, internal review and re-curation of publicly available curations led to the inclusion of 63 gene-disease pairs, representing 29% of the gene-disease pairs on the final panel (**Figure 2**).

**Figure 2.**
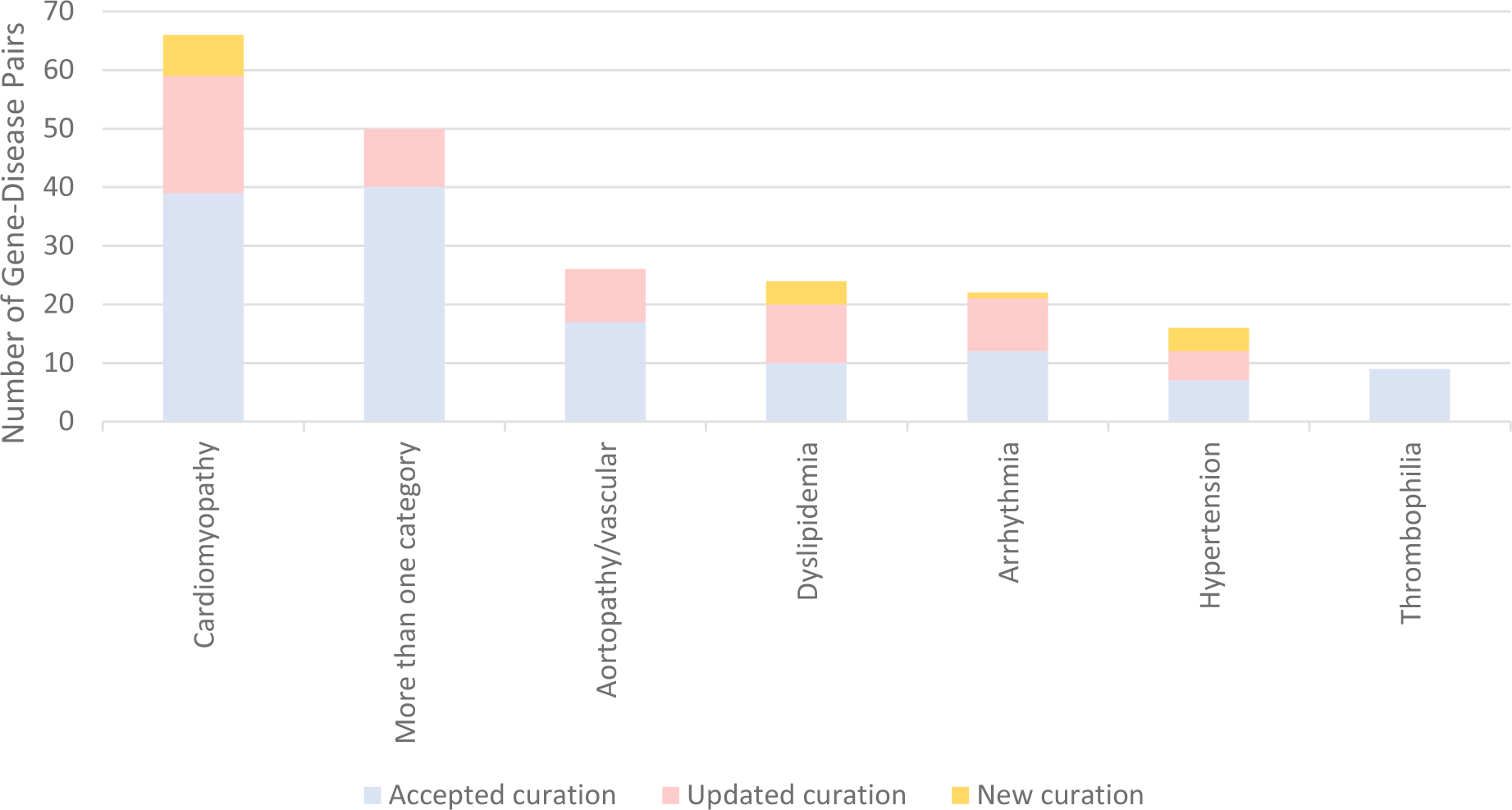
Number of Included Gene-Disease Pairs in Each CVD Phenotype by Curation Source. All included gene-disease pairs had a GDR classification of strong or definitive based on the ClinGen Gene-Disease Validity Framework. Included gene-disease pairs came from one of three sources: 1) an accepted publicly available or internal curation, 2) an updated publicly available or internal curation or 3) a new internal curation. Updated curations and curations of gene-disease pairs with no current publicly available or internal curation data led to the inclusion of 79 gene-disease pairs on the total 215 gene-disease pair CVD panel.

To support test scalability and to prioritize the return of medically actionable information, variants identified in genes on the CVD gene panel were required to meet a pathogenic or likely pathogenic (P/LP) variant classification threshold for return, consistent with ClinGen guidance supporting return of P/LP variants in gene-disease pairs with a D/S classification^31^.

#### Secondary Findings Gene Panel

The American College of Medical Genetics and Genomics (ACMG) recommends testing for a minimum list of gene-disease pairs deemed to be medically actionable in the context of genomic sequencing-based diagnostic tests^32^. These non-diagnostic results, or ‘secondary findings’, include genes associated with inherited cancer risk, CVD, and metabolic conditions. In alignment with ACMG recommendations, analysis of secondary findings genes on the current version of the ACMG list is included as a component of the TruGenome^TM^ CVD Test. Of the 81 current genes recommended for evaluation, 46 genes were included in the CVD panel, and the remaining 35 non-CVD genes were incorporated for testing as secondary findings. Variants identified in these 35 genes required a classification of P/LP in alignment with reporting of the CVD panel and ACMG recommendations.

#### Cardiovascular Disease Risk Alleles

A risk allele is defined as a genetic variant, which may or may not be common in the population, that is associated with an increased probability of an individual developing a disease when compared to the baseline risk observed in a control population without the allele^19^. The evaluation of CVD-associated risk alleles was framed using recommendations from the ClinGen Low Penetrance/Risk Allele Working group^19^. The risk allele reporting threshold included classification as an Established Risk Allele or an Established Risk Genotype. Relevant risk alleles or genotypes in six genes were identified through a comprehensive literature review, assessment of risk alleles previously evaluated in ICSL in support of a rare undiagnosed disease genome test, and through consultation with medical genetics experts. Two risk alleles and two risk genotypes met criteria for reporting: the Factor V Leiden variant in *F5,* the prothrombin variant in *F2,* the E2/E2 genotype in *APOE* and homozygosity or compound heterozygosity for the G1 and G2 alleles in *APOL1.* (**Table 1**). Heterozygosity for the E2 or E4 allele in *APOE* and heterozygosity of the c.253G>A p.(Asp85Asn) variant in *KCNE1* were evaluated and did not meet the threshold for reporting based on the absence of replicated case-control studies supporting an associated >2-fold increased risk of CVD and long-QT syndrome, respectively.

**Table 1.** Cardiovascular Disease Risk Alleles.

| Gene | Allele/Genotype | Associated Risk | Classification | Outcome | Supporting Evidence |
| --- | --- | --- | --- | --- | --- |
| *F5* | Heterozygosity or homozygosity for c.1601G>A (p.Arg534Gln) | Thrombophilia susceptibility | Established Risk Allele | Yes | Odds ratio >2 for increased risk for thromboembolism<br>Replication across studies |
| *F2* | Heterozygosity or homozygosity for c.*97G>A variant | Thrombophilia susceptibility | Established Risk Allele | Yes | Odds ratio >2 for increased risk for thromboembolism<br>Replication across studies |
| *APOE* | Homozygosity for E2 allele [rs429358 (T:T) and rs7412 (T:T)] | Familial dysbetalipoproteinemia | Established Risk Genotype | Yes | Odds ratio >2 for increased risk for cardiovascular disease<br>Replication across studies |
| *APOL1* | Homozygosity or compound heterozygosity for G1 and G2 alleles | Chronic kidney disease | Established Risk Genotype | Yes | Odds ratio >2 for increased risk for chronic kidney disease with associated hypertension<br>Replication across studies |

#### Clinical Pharmacogenomic Report

PGx variants endorsed by the FDA to have a potential impact on therapeutic management recommendations, safety or response, or pharmacokinetic properties^20^ and with published Clinical Pharmacogenetics Implementation Consortium (CPIC) guidelines^21^ as of May 2022, were reviewed for evidence of clinical and functional impact. Guidance from the Association for Medical Pathology (AMP) on allele selection was also reviewed^22^. Variants in ten genes associated with a total of 38 drugs were validated for inclusion (**Table 2**). Variants in *CYP2C19, CYP2C9, CYP4F2, SLCO1B1,* and *VKORC1* are associated with drugs with implications for the management of CVD conditions. The remaining genes are associated with drugs that may be prescribed to individuals with cardiovascular events, for example, immunosuppressants following organ transplantation, antidepressants, and pain medications. Chemotherapy, HIV treatment, and proton pump inhibitors were also included. PGx reporting follows ACMG recommendations^33^.

**Table 2.** Gene-Drug Pairs Included in Pharmacogenetic Testing.

| **Gene** | **Associated Drug(s)** | **Variant/Star Allele(s) Included** |
| --- | --- | --- |
| ***CYP2C19*** | Citalopram, Clopidogrel, Doxepin, Escitalopram, Lansoprazole, Omeprazole, Pantoprazole, Voriconazole | *2, *3, *4.001, *4.002, *5, *6, *7, *8, *9, *10, *17, *35 |
| ***CYP2D6*** | Amitriptyline, Atomoxetine, Clomipramine, Codeine, Desipramine, Doxepin, Imipramine, Fluvoxamine, Nortriptyline, Paroxetine, Tamoxifen, Tramadol, Trimipramine | *2, *3, *4.013, *4, *5, *6, *7, *8, *9, *10, *12, *13, *14, *15, *17, *21, *29, *31, *36, *40, *41, *42, *49, *56, *59, *68, copy number variants, tandem and hybrid variants |
| ***CYP3A5*** | Tacrolimus | *3, *6, *7 |
| ***CYP2C9*** | Celecoxib, Flurbiprofen, Fosphenytoin, Ibuprofen, Meloxicam, Phenytoin, Piroxicam, Warfarin | *2, *3, *5, *6, *8, *11, *12, *13, *14, *15 |
| ***VKORC1*** | Warfarin | c.-1639G>A (rs9923231) |
| ***CYP4F2*** | Warfarin | *3 |
| ***DPYD*** | Capecitabine, Fluorouracil | c.[1236G>A;1129-5923C>G] (rs56038477; rs75017182), c.1679T>G (rs55886062), c.1905+1G>A (rs3918290), c.2846A>T (rs67376798), c.557A>G (rs115232898) |
| ***HLA-B***<br>**(\*57:01**<br>**proxy: *HCP5*)** | Abacavir | n.852T>G (rs2395029) |
| ***SLCO1B1*** | Simvastatin, Atorvastatin, Rosuvastatin | *5 |
| ***TPMT*** | Azathioprine, Mercaptopurine, Thioguanine | *2, *3A, *3B, *3C, *4 |

#### Polygenic Risk Score for Coronary Artery Disease

Comprehensive review of available PRSs, and the evidence supporting their validity and potential clinical utility, supported inclusion of a CAD PRS into the test design (see Supplemental Methods). After comparing the performance metrics and integration feasibility, an ancestry specific CAD PRS developed by Allelica, Inc. was selected for use^35^. A bespoke CAD PRS test report was developed which reports a binary ‘elevated’ or ‘not elevated’ outcome. The elevated threshold was defined as an estimate that the tested individual is at 2-fold increased risk of developing CAD compared to the remainder of the individuals in their ancestry group. The upper and lower bounds of the confidence interval are also reported to highlight uncertainty in the threshold estimate. Language regarding uncertainty of PRS predictions were included and the importance of genetic variants which may not be included in the PRS calculation, medical history, family history, and lifestyle factors was highlighted (**Supplemental Figure 3**). Given the uncertainty of the clinical utility of PRS scores the TruGenome^TM^ CVD Test CAD PRS is provided as Research Use Only (RUO) finding, and the associated report highlights that the result is not validated for use for the diagnosis, prevention, or treatment of CAD.

### Evaluation of test performance in the 1000 Genomes Project cohort

To ascertain TruGenome™ CVD Test performance, the frequency of findings across the monogenic CVD genes, risk allele, and PGx components was evaluated in 2,594 unrelated individuals from the 1,000 Genomes Project (1KGP) cohort. PRS performance was not assessed as the 1000 Genomes Project data was used in the development of the Allelica multi-ancestry CAD score^25^. For this analysis, likely reportable variants were defined as rare variants (MAF <1%) in genes on the CVD and/or secondary findings gene panels that have a P/LP ClinVar 2* classification or were predicted to have the functional effect of protein truncation (and thus have a high probability of being classified as LP/P) for which this variant type is an established disease-causing mechanism.

One hundred and thirty-five genomes (6%) had a single likely reportable variant (n= 131 genomes) or two likely reportable variants (n=4 genomes) identified in the nuclear genes associated with autosomal dominant or semi-dominant conditions on the CVD gene panel. No genomes had two likely reportable variants/alleles in a gene associated with autosomal recessive disease. The common *TTR* c.239C>T p.(Thr80Ile) variant associated with hereditary transthyretin amyloidosis was identified in 28 genomes (1%). Thirty-seven (35%) of the 105 unique variants were P/LP ClinVar 2* variants and 68 (65%) were predicted truncating variants. Likely reportable variants were identified most often in African populations, which was driven by *TTR* c.239C>T, p.(424G>A) (n=20) and predicted truncating variants (n=17).

Twenty-four genomes (0.9%) had a single likely reportable variant in one of the non-CVD secondary findings genes associated with autosomal dominant or semi-dominant conditions. These findings included 22 unique variants, 83% of which were P/LP ClinVar 2*. Secondary findings were identified in 17 of the 27 1000 Genomes Project population groups, with Northern and Western European (n=6) and African (n=7) populations accounting for nearly all these findings.

A risk allele was identified in 159 genomes (6%) (**Supplemental Table 3**). The most frequently identified risk alleles were homozygosity or compound heterozygosity for the G1 and G2 alleles in *APOL1* (4% of genomes) and heterozygosity for the factor V Leiden variant in *F5* (1% of genomes).

Almost all genomes (2,586, >99%) had at least one PGx finding reported. Each genome had on average of two PGx findings (range 0-5) in genes associated with a drug that has direct CVD implications, and two PGx findings (range 0-5) in the remaining genes on the pharmacogenomic test. Overall, 93% of genomes had one or more PGx finding in a gene associated with a drug with direct CVD implications (**Figure 3 and Supplemental Table 3**).

**Figure 3.**
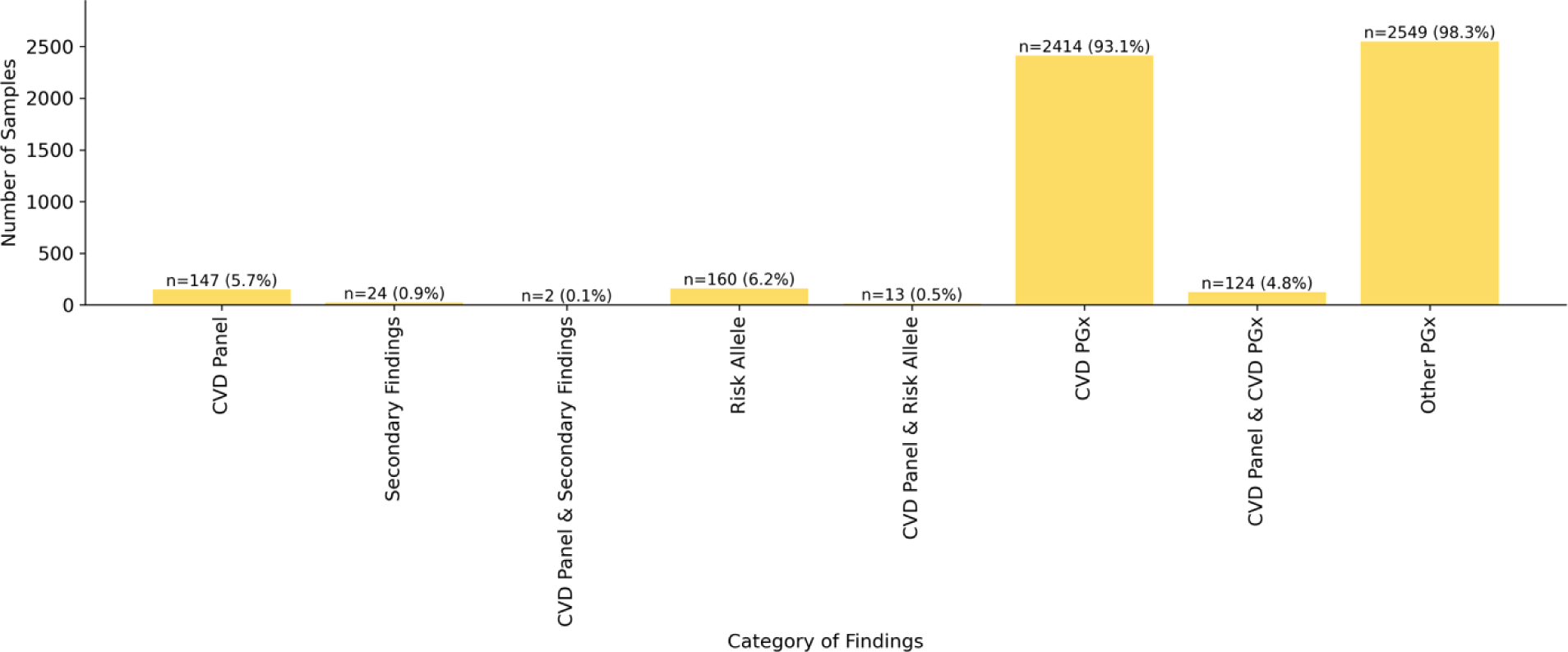
Percent of Genomes from an Unselected Population with Genetic Disease, Genetic Risk and PGx Findings in the TruGenome^TM^ CVD Test. To ascertain TruGenome^TM^ CVD Test performance, the frequency of findings across the monogenic disease, risk allele, and PGx components was evaluated in 2,594 unrelated individuals in the 1,000 Genomes Project cohort. Six percent of genomes had a finding in a CVD-associated gene, 6% had a risk allele finding, 0.9% had a non-CVD-associated ACMG secondary finding, and 93% with at least one CVD-associated PGx variant.

### TruGenome^TM^ CVD test performance compared to prior genetic testing

TruGenome^TM^ CVD performance was assessed in a blinded cohort of 20 individuals with a clinical diagnosis of a CVD who had previously received genetic testing (**Table 3**). Most individuals (16/20, 80%) were between the ages of 40-60 years old and approximately half had cardiomyopathy as a component of their CVD phenotype (11/20, 55%).

**Table 3.** Age, Phenotypes, and Clinical Testing Details for Samples from Individuals at Risk for Genetic CVD.

| Variable | Number of Individuals |
| --- | --- |
| Age (years) |  |
| 20-40 | 2 |
| 40-60 | 16 |
| >60 | 2 |
| Phenotype(s) |  |
| Aortopathy/aneurysms | 5 |
| Arrhythmia only | 1 |
| Cardiomyopathy and arrhythmia | 3 |
| Cardiomyopathy and/or heart failure | 8 |
| Hyperlipidemia | 3 |
| Clinical genetic testing |  |
| Aortic dysfunction/dilation panel | 5 |
| Cardiomyopathy panel | 7 |
| Comprehensive cardiac panel | 5 |
| Familial hypercholesterolemia panel | 2 |
| Long QT syndrome panel | 1 |

The average number of variants triaged in genes on the CVD panel and in non-CVD secondary findings genes per case was four (range 1 to 10 variants). Of the 77 variants triaged across the 20 cases, most were single nucleotide variants (SNVs; 67/77, 87%) and in genes on the CVD gene panel (59/77, 77%) vs. in non-CVD secondary finding genes.

Time to triage, interpret and report each case, including laboratory director review and sign out, ranged from 9 to 96 minutes. For cases that did not require a curation and without a P/LP variant reported (n=12) the average time was 20 minutes, ranging from 9 to 48 minutes. For those with a P/LP variant that had been previously curated, the range was 19 to 43 minutes (n=3). The range was 58 to 96 minutes for cases with one or more reportable P/LP variants requiring curation (n=5).

Seven P/LP variants were identified in genes on the CVD panel (*FBN1, MYBPC3* x3*, SERPINC1, TTN* x2) and one P/LP variant was identified in a non-CVD secondary findings gene (*RYR1*) (**Table 4**). One or more pharmacogenomic variant in genes associated with drugs determined to have direct CVD implications were identified in all 20 individuals.

**Table 4.** Findings in Individuals with a Suspected Diagnosis of Genetic CVD.

| Category of Result | Individuals with a Reportable Finding<br>N, (% of Total) |
| --- | --- |
| **CVD gene panel** | 7, (35) |
| **CVD risk allele** |  |
| *F5* | 0, (0) |
| *F2* | 1, (5)* |
| *APOE* | 0, (0) |
| *APOL1* | 1, (5)** |
| **Non-CVD ACMG secondary findings** | 1, (5) |
| **Elevated CAD PRS** | 0, (0) |
| **Pharmacogenomic finding(s)** |  |
| **CVD-related** | 20, (100) |
| **Non-CVD related** | 16, (80) |

\*Heterozygosity for the c.\*97G>A risk allele variant in F2 was identified
\*\* *The APOL1* genotype G1/G2 was identified

All P/LP variants in the CVD gene panel identified on the TruGenome^TM^ CVD test had been previously identified and reported by the clinical genetic testing performed by Greenwood Genetic Center, except a P variant in the *SERPINC1* gene which was only detected by TruGenome^TM^ CVD test. Variants in *SERPINC1* are associated with antithrombin III deficiency. This individual had a Comprehensive Cardiac NGS Panel clinical test at Greenwood Genetic Center based on a diagnosis of obstructive cardiomyopathy and *SERPINC1* was not part of the panel of genes tested.

## Discussion

Genetic testing for patients with CVD is substantially underutilized, despite guidelines and established evidence of benefit. While many factors impact access to testing, the development of a comprehensive assay that enables concurrent assessment of multiple CVD-associated genetic factors on a genome backbone may support both laboratory and clinician adoption. This approach reduces the risk of false negative results that can occur when using focused NGS gene panel testing, enables the evaluation of complex CVD presentations which may have multiple genetic drivers, and supports the detection of variants that may not be assessed with other approaches. A broad testing approach also provides the option to identify ancillary genetic risk information including medically actionable secondary findings and pharmacogenomic information. A GS-based CVD genetic risk assessment may address genetic testing inconsistencies, underutilization and underdiagnosis in CVD by providing a single, scalable, multifaceted molecular diagnostic platform.

Gene curation is a necessary, and sometimes overlooked, step in robust clinical test development. The level of evidence supporting the association between a gene-disease pair impacts whether an identified variant in a gene would be expected to increase risk for the associated disease. The gene curation efforts described here expanded the CVD gene panel from 139 gene-disease pairs with previously curated D/S classifications to a total of 215 gene-disease pairs. Clinical laboratories, population genomic studies, and other institutions performing genomic analysis for individuals with CVD may consider utilizing this panel as a resource. Additions and subtractions are expected to be necessary as new information becomes available about these and other genes.

Polygenic risk scores have shown utility for population-based risk stratification, but their applicability to clinical medicine as individual level risk estimates across diverse ancestry groups is under investigation^34^. Data from a recent studies supports benefits and changes in patient management with the return of combined CAD PRS and monogenic disease risk information^35^.

However, more evidence is required to understand the clinical utility of the application of PRSs across ancestry groups and to inform best practices for PRS reporting to patients and providers. This ambiguity was a factor in our decision to return the CAD PRS finding as an RUO finding. Evidence generated from clinical trials integrating CAD PRSs as a component of a comprehensive genome-based test may help to fill these gaps and inform future integration of PRSs into clinical practice.

While the intended use of the TruGenome^TM^ CVD Test is for adult patients with a diagnosis of CVD that may be genetic, the test development approach described here can inform the development and implementation of GS-based CVD risk assessment tests for broader populations. The rate of 6% of genomes in a large, unselected population having one or more likely reportable variant in the CVD gene panel aligns with expectations based on disease prevalence and penetrance^36^. Based on our test performance evaluation in a small cohort of individuals with CVD, the average number of variants requiring triage per case was 4, and the average time for interpretation and reporting for cases without a P/LP variant was 20 minutes. These data suggest that the TruGenome^TM^ CVD Test and similar broad GS based CVD-risk assessment tests are likely amenable for population scale implementation, given that most individuals would not have a P/LP variant identified and therefore would not require time intensive interpretation and reporting.

This work has several limitations. The CVD gene panel content reported here may not represent all relevant gene-disease pairs with a D/S association, as available evidence to evaluate GDRs is continually generated. Similarly, there are likely other CVD risk alleles that could meet the threshold for inclusion that were either not evaluated or did not have sufficient evidence to support inclusion at the time of test development. The rate of genomes with a PGx finding in a gene-drug pair with direct CVD implications was not interpreted in the context of the dosing algorithm for warfarin, which incorporates the alleles tested in *CYP2C9, VKORC1* and *CYP4F2* in combination to modify warfarin dosing ^37^. The data from our evaluation of test performance in a cohort of individuals suspected to have genetic CVD was based on a small number of cases and may not be generalizable to other laboratory workflows. Finally, time estimates in the test performance evaluation were based on the use of internally developed software for the reporting of risk alleles, the CAD PRS and PGx findings without manual data review.

GS as a platform for CVD genetic risk assessment in adult patients with CVD phenotypes for which genetic testing is recommended may increase access to relevant and actionable genetic risk information. This approach is scalable, transferrable to other disease areas, and supports the use of genetic data across the lifespan of an individual.

## Data Availability

All data produced in the present study are available upon reasonable request to the authors.

## Non-Standard Abbreviations and Acronyms

ACMG: American College of Medical Genetics and Genomics
AMP: Association for Molecular Pathology
CVD: cardiovascular disease
GDR: gene-disease relationship
ICSL: Illumina Clinical Services Laboratory
MAF: minor allele frequency
GS: genome sequencing
P: pathogenic
LP: likely pathogenic

## Acknowledgments

We thank the ICSL Interpretation and Reporting Team for their support and feedback during test development work.

## Sources of Funding

This work was supported by Illumina Inc. David E. Lanfear’s effort is supported in part by grants from the NIH (R01HL132154; P50MD017351).

## Disclosures

LMA, AJC, JL, JA, AM, AC, JPT, RR, CMB, KGG, RH, PM, BM, FM, AW, AH-V, ML, DL, DH, SSA, AK, SPS, EM and RJT were employees and/or shareholders of Illumina, Inc. when this work was completed. DEL is a consultant for Abbott Laboratories, ARMGO, Astra Zeneca, Illumina, Janssen, and Martin Pharmaceuticals, and has participated in clinical research from Akros, AstraZeneca, Bayer, Illumina, Janssen, Lilly, and Pfizer, and has a patent (held by Henry Ford Health) for a beta blocker polygenic score.

## Supplemental Material

Supplemental Methods

Tables S1-S3

Figures S1-S3.

## Notes

### Author Declarations

Institutional review board approval for activities involving human samples was received from WIRB-Copernicus Group (Protocol 20241866). Informed consent was not required.

