## Supplemental Materials for "Development of a comprehensive cardiovascular disease genetic risk assessment test"

#### Supplemental Methods

##### *Cardiovascular Disease Gene Panel Development*

Conditions and phenotypes relevant to adults with CVD in a cardiac clinic setting were considered for inclusion in the CVD gene panel. Congenital heart defects (CHDs) were not assessed based on the complexity of genomic contributions to CHD and their likely presentation in childhood. Cerebrovascular disease was also not assessed based on the likely initial neurologic presentation of these conditions. Aortopathy, arrhythmia, cardiomyopathy, and dyslipidemia were included as relevant phenotypes based on recommendations for genetic testing for adults with one of these diagnoses [9,10]. Thrombophilia and hypertension were also considered relevant phenotypes based on the established link with associated genetic risk and potential implications for medical management in a cardiac clinic setting.

Genes associated with aortopathy, arrhythmia, cardiomyopathy, dyslipidemia, hypertension, or thrombophilia were identified through comprehensive literature review, evaluation of publicly available and internal gene-disease curations, and assessment of commercially available CVD gene panels. Nuclear DNA and mitochondrial DNA genes associated with primary mitochondrial disease, which can include CVD phenotypes and present in adulthood, were identified from the Mitochondrial Disease Compendium[11] with additional review of the literature.

For monogenic conditions, gene-disease pairs were evaluated for both the quantity and quality of evidence supporting the purported gene-disease relationship (GDR). A modified version of the ClinGen Gene-Disease Validity framework was utilized and a minimum classification of definitive or strong (D/S) was implemented as the threshold for inclusion of a gene-disease pair. Briefly, for each GDR, the ClinGen framework awards points for both genetic and experimental evidence supporting the relationship and assigns a classification to the GDR based on a weighted matrix. Genetic evidence is awarded a maximum of 12 points and experimental evidence a maximum of 6 points. To reach a classification of D/S, the total score from both genetic and experimental evidence must reach 12 points. If there is not sufficient genetic evidence to reach 6 points, even with the maximum score for experimental evidence, the final classification cannot reach D/S. Scoring of evidence for GDR curation of mitochondrial DNA genes in association with primary mitochondrial disease was based on internal expertise and guidance from the ClinGen Mitochondrial Diseases Gene Curation Expert Panel (GCEP). Gene curation took place between October 2021 and July 2022 and gene panel inclusion decisions were made based on information available at the time of curation.

Gene-disease pairs with D/S gene-disease relationship (GDR) classifications from prior internal Illumina Clinical Services Laboratory (ICSL) gene curations, ClinGen, or groups that use the ClinGen framework were included in the CVD gene panel. Gene-disease pairs with a refuted or disputed relationship per internal curations, ClinGen, or groups using the ClinGen framework were not included. All remaining nuclear DNA gene-disease pairs, including those with no publicly available GDR classification, or with a moderate or lower GDR classification from any group, were curated using a modified version of the ClinGen Gene-Disease Validity framework[12] **(Supplemental Figure 1)**. The evaluation of evidence was dependent on the classification of the GDR (i.e. definitive, strong, moderate etc.,) and whether the GDR source was from 1) ClinGen, 2) a group that utilizes the ClinGen framework (including ICSL), 3) a group that uses an approach based on the ClinGen framework, or 4) a group that uses an alternate methodology. GDRs with a D/S classification from a group using a ClinGen framework-based approach were verified via assessment by two clinical scientists to evaluate if the evidence cited for the GDR met the classification criteria. A literature search was performed for select GDRs to assess if new evidence warranted an update to the available curation. GDRs deemed to have sufficient new evidence in the literature to potentially change the GDR classification were assessed using an expedited curation process, which consisted of an initial assessment of genetic evidence supporting the GDR. Experimental evidence was only evaluated for gene-disease pairs with sufficient genetic evidence scores (≥6 points) to potentially be classified as D/S. *DMPK* and *CNBP*, associated with and Myotonic Dystrophy type 1 and type 2, respectively, were included without gene curation given the established association between these genes and myotonic dystrophy, which can include cardiac arrhythmias and cardiomyopathies as part of the phenotypic spectrum.

##### *Removal of Common Variants in CVD and non-CVD Secondary Finding Genes from Case Analysis*

To reduce variant triage burden during interpretation, common variants in the CVD ene panel and non-CVD secondary findings genes that were unlikely to be disease causing given the penetrance and prevalence of the associated disease were proactively excluded from case analysis. Disease prevalence and average penetrance were estimated for each of the associated diseases as follows: disease prevalence and average penetrance were collected from the literature, GeneReviews[13] and Orphanet[14] for each of the associated diseases. When specific penetrance estimates were not available from the literature, a permissive default penetrance of 80% and 40% was assigned to autosomal recessive and autosomal dominant conditions, respectively [15]. Similarly, conditions without available disease prevalence information were assigned a default prevalence of 1/250,000 to avoid removing variants inappropriately. This prevalence cutoff was informed by definitions of rare disease based on prevalence from the US Rare Disease Act[16] and the European Union [17]. Next, allele count (AC) and allele number (AN), defined as the allele count in genotypes for each alternate allele and the total number of alleles in called genotypes, respectively, were extracted from gnomAD v2.1.1 [18] for variants that were likely to impact the protein sequence and were present with an overall minor allele frequency (MAF) <5%. MAF population cutoffs were then calculated using the below equations for diseases with autosomal dominant or X-linked and autosomal recessive inheritance, respectively.

$$AD\ or\ X-linked\ autoscore = \mathrm{Log}_{10}\frac{\left[\left(1-\left(1-95\%\ \mathrm{CI\ lower\ bound\ of}\ \frac{\mathrm{AC}}{\mathrm{AN}}\right)^2\right)*\mathrm{Penetrance}\right]}{\mathrm{Prevalence}}$$

$$AR\ autoscore = \mathrm{Log}_{10}\frac{\left[\left(95\%\ \mathrm{CI\ lower\ bound\ of}\ \frac{\mathrm{AC}}{\mathrm{AN}}\right)^2*\mathrm{Penetrance}\right]}{\mathrm{Prevalence}}$$

Variants with a score >0 were deemed too common to be P/LP based on the estimated disease prevalence and penetrance and were removed from the interpretation and reporting workflow for case analysis. Of a total 1,666,138 unique variants from gnomAD v2.1.1 in the CVD panel and non-CVD ACMG secondary findings genes, 428,380 variants were removed with an average number of variants removed per gene of 2,532 (range 41 to 60,826 variants).

##### *Evaluation of Cardiovascular Disease Risk Alleles*

Relevant risk alleles for adult cardiac clinic patients were identified through literature review, prior internal laboratory experience and consultation with medical genetics experts. Evaluation of risk alleles was based on guidance from the ClinGen Low Penetrance/Risk Allele Working group [19]. Each risk allele was evaluated with respect to the size of case-control cohorts reported in the literature, replication of the association between the risk allele and disease phenotype across studies, and any potential bias. The odds ratio point estimate of a minimum >2-fold increased risk of disease was set as the threshold for including a risk allele, and the terminology of Established Risk Allele, Likely Risk Allele and Uncertain Risk Allele was adopted from the ClinGen Low Penetrance/Risk Allele Working group guidance to classify the level of evidence supporting each evaluated association. Associations for which a genotype that combines more than one risk allele is associated with increased risk for disease were categorized and labeled as Established Risk Genotypes, Likely Risk Genotypes and Uncertain Risk Genotypes.

##### *Identifying and Evaluating Gene-Drug Pairs for Pharmacogenomic Testing*

Gene-drug pairs for pharmacogenomic testing were selected based on guidance from the US Food and Drug Administration (FDA), the Clinical Pharmacogenetics Implementation Consortium (CPIC) and the Association for Molecular Pathology (AMP). Guidance from the ACMG informed the reporting strategy. Genes and drugs present in the FDA table of pharmacogenomic associations [20] and with published CPIC guidelines [21] as of May 2022 were identified. Variants in genes with sufficient evidence for a clinical and functional impact were selected for the test. These include tier 1 (recommended) and tier 2 (optional) variants from AMP publications[22], together with variants with sufficient literature evidence reviewed and confirmed by CPIC. Variants with uncertain clinical impact or function were excluded. Gene-drug pairs were identified as being related to CVD based on review of the indications for prescribing the associated drugs (as defined by CPIC and the FDA) and were confirmed through consultation with a subject matter expert in CVD pharmacogenomics (DL).

##### *Evaluating Polygenic Risk Scores for Cardiovascular Disease*

Polygenic risk scores (PRSs) for CVD phenotypes, including CAD and atrial fibrillation, were investigated for potential inclusion in the test. Evidence on the extent and robustness of the PRS validation, strength of risk reclassification and clinical utility were assessed. For each publication describing a CVD-related PRS the following were collated: size and diversity of the population tested, the number of markers used in the PRS calculation and the performance characteristics of the score. CAD was determined to be the CVD phenotype with the most robust evidence supporting integration into clinical testing.

Three CAD PRS scores were evaluated for inclusion including the GPS CAD [23], metaGRS [24] and an ancestry specific Allelica PRS score[25]. The latter was chosen based on its overall performance metrics and integration feasibility given the availability of a validated Docker image. A detailed description of the development and validation of the Allelica, Inc. ancestry-specific CAD PRS is available elsewhere[25]. Briefly, the Allelica, Inc. CAD PRS calculates genetic ancestry for each individual and a corresponding ancestry specific PRS is applied. Computed ancestry groups include African, American Admixed, East Asian, European, and South Asian. Elevated risk of CAD is defined as an individual with $\geq$2-fold increased relative risk of developing CAD compared to the remainder of the individuals in their ancestry group. The proportion of individuals in each ancestry group with a $\geq$2-fold increased risk ranges from 0.22 to 0.17 across computed ancestry groups. The OR per SD for the 5 ancestry-specific PRSs ranges from 1.323 to 1.579 and the range of AUROCs was 0.69 to 0.78.

To inform PRS reporting we reviewed the literature for best practices for the return of genetic risk information from a PRS[26]. We carefully considered how to convey the uncertainty of reporting the PRS as population-level risk stratification in the context of a test focused on reporting patient specific risk information. We also considered how to convey the current state of evidence regarding the use of PRSs for clinical decision making with respect to disease risk assessment and management.

##### *Assessment of CVD-related findings in the 1000 Genomes Project cohort*

Whole genome sequence data from a total of 2,594 unrelated 1000 Genomes Project individuals were analyzed. Small variants from the NYGC BWA/GATK joint call dataset[27] were used to assess the number and burden of reportable CVD gene panel findings and non-CVD secondary findings. Small variants,

CNVs, and gene-specific callers from DRAGEN v4.03 individual analyses[28] were used to assess risk alleles and pharmacogenomic findings. Variants in the mitochondrial genome were excluded from this analysis given the absence of information on heteroplasmy levels.

Potentially relevant monogenic variants were annotated using Nirvana and filtered such that only those with a MAF <1% and either classified as P/LP in ClinVar with a 2* status or with the predicted functional effect of protein truncation were included in the dataset. Protein truncation variants were defined as those that resulted in a frameshift, canonical splice disrupting, and stop-gain in genes where loss of function (LOF) is an established mechanism of disease. The determination of LOF as a disease mechanism was based on review of ClinGen resources (e.g., haploinsufficiency scores, GDR curation summary statements, applicable Variant Curation Expert Panel (VCEP) guidelines) and previously reported internally classified P/LP LOF variants in relevant genes [29]. Protein truncating variants were reviewed to confirm they would not be expected to escape nonsense mediated decay (NMD) based on their position within the protein. The MANE select transcript was chosen as the clinically relevant transcript, except in those cases where a gene had more than one MANE transcript. For these genes, the clinically relevant transcript was chosen based on consensus use in ClinVar, HGMD, and MANE. In the case where there was no consensus, a transcript was chosen based on expression pattern, literature review, and protein length, such that the longest isoform expressed in the tissue of interest was preferred. (Selected transcripts are available upon request.) Truncating variants in *TTN* were included only if they were in exons that are highly expressed in cardiac tissue (defined as exons with a percent spliced in (PSI) of >90%) based on the established association with dilated cardiomyopathy [30]. Variants identified in more than one genome were manually reviewed to confirm their inclusion as likely reportable variants. Genomes with more than one variant identified in the same gene were also manually reviewed to rule out the presence of a single complex heterozygous allele.

The number of genomes in the dataset with one or more variants in genes associated with autosomal dominant or semi-dominant conditions, the number of genomes with two variants in genes associated with autosomal recessive conditions, the number of unique likely reportable variants and the proportion of truncating variants that were not in ClinVar as 2* P/LP, and the number of likely reportable variants across ancestry groups were calculated.

CVD risk allele status was also determined for each genome in the dataset. For each genome, the DRAGEN v4.0.3 small variant and CNV gVCFs were filtered to the seven risk allele sites in the TruGenome™ CVD test. For each site, genotypes were mapped to a predefined matrix to determine the presence or absence of risk alleles.

Finally, a pharmacogenomic analysis was conducted for each genome. Pharmacogenomic star alleles were called using the DRAGEN v4.0.3 small variant & CNV output. Star alleles were mapped to a matrix of functional or numerical scores to determine the impact for each drug-gene pair.

##### *Evaluating Performance with Previously Tested Clinical Samples*

To evaluate overall test performance, DNA from 20 individuals with a suspected hereditary CVD were tested via the TruGenome™ CVD Test. Individuals with indications for testing that aligned with the intended use population for the test were selected from a cohort of previously tested patients at Greenwood Genetics Laboratory (GGL). Samples from individuals in the GGL cohort with a P/LP variant identified via clinical testing were prioritized for inclusion. ICSL was blinded to prior test outcomes until after the completion of the TruGenome™ CVD analysis.

Variants in the CVD gene panel and non-CVD secondary findings genes were triaged and interpreted by 2 PhD clinical scientists and 1 genetic counselor, who tracked the number of variants requiring triage per case and time for interpretation, review, and reporting. Variant curations were only performed for variants determined by the analyst to have a high likelihood of being classified as P/LP and reported. Only cases with P/LP classified variants in the CVD and/or non-CVD ACMG SF gene panels were reviewed by the genetic counselor. Interpretations and classifications were reviewed by a laboratory director (AK) who also tracked the total time engaged in test review and reporting. The presence of risk alleles, the PRS outcome (elevated or not elevated) and the presence of PGx variants were autogenerated by the analysis pipeline using two internally developed software services integrated with the variant analysis system, TruSight Software Suite.

The number of individuals with a finding in the CVD gene panel, select CVD risk alleles, non-CVD secondary findings, elevated CAD PRS, and both CVD- and non-CVD-related pharmacogenomic information was calculated.

### Supplemental Tables

#### Supplemental Table 1.

Risk alleles identified in unrelated individuals in the 1000 Genomes Project dataset.

| **Risk Allele** | **Number of Genomes (N, %)** |
|---|---|
| *APOL1* | |
| Homozygosity G1/G1 | 57, 2% |
| Homozygosity for G2/G2 | 10, <1% |
| Compound heterozygosity for G1/G2 | 44, 2% |
| *F5* | |
| Heterozygosity for c.1601G>A (p.Arg534Gln) | 29, 1% |
| Homozygosity for c.1601G>A (p.Arg534Gln) | 1, <1% |
| *F2* | |
| Heterozygosity for c.*97G>A | 17, 1% |
| Homozygosity for c.*97G>A | 1, <1% |
| *APOE* | |
| Homozygosity for E2 allele [rs429358 (T:T) and rs7412 (T:T)] | 0, 0% |

[Please see supplemental Excel spreadsheet]

#### Supplemental Table 2.

**CVD Gene panel Gene Disease Pairs.** The CVD gene panel includes 215 gene-disease pairs that have a strong or definitive (S/D) gene-disease relationship (GDR). S/D GDRs were accepted from groups that use the ClinGen Gene-Disease Validity framework for gene curation. All other gene-disease pairs were either curated internally using an expedited approach or verified. The number of genetic evidence points, experimental evidence points and the references supporting each GDR are presented in Supplemental Table 2.

[Please see supplemental Excel spreadsheet]

#### Supplemental Table 3.

**TruGenomeCVD$^{TM}$ Test Findings in the 1000 Genomes Project Across the 2,594 Genomes.** The number of findings in each of the 2,594 genomes in the CVD gene panel, non-CVD ACMG secondary findings genes, risk alleles and CVD-related and non-CVD-related PGx alleles is presented in Supplemental Table 3. The details for each finding are also presented across these four components.

#### Supplemental Figures

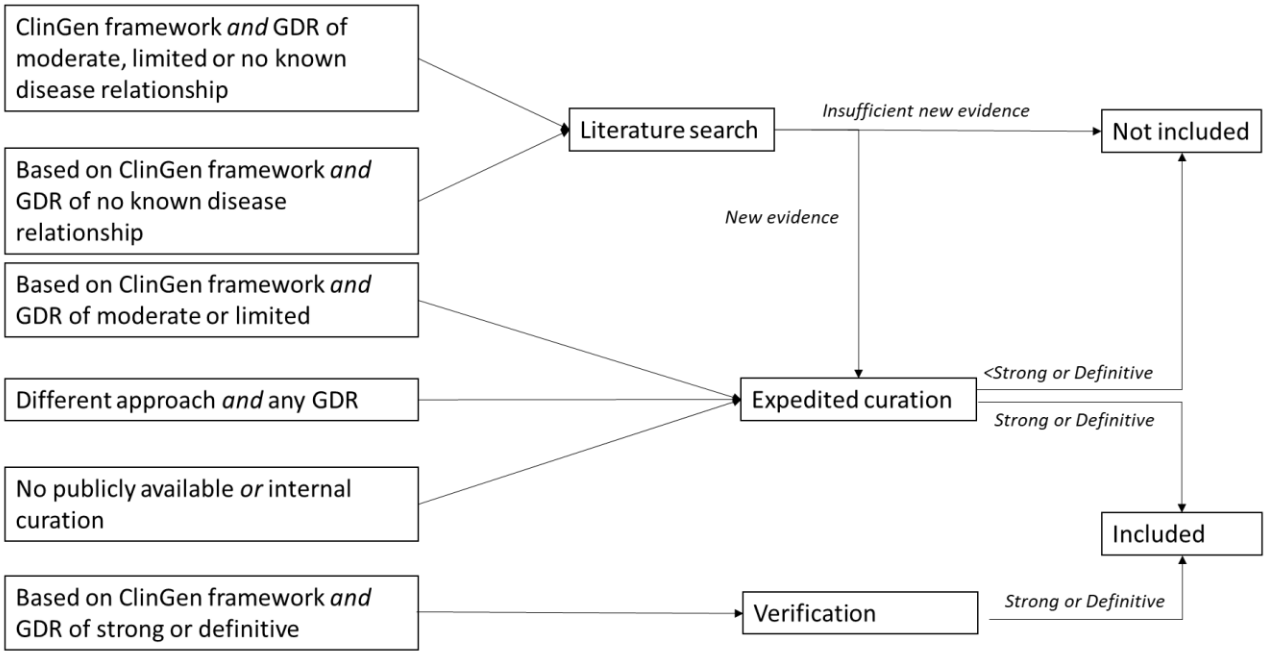

##### Supplemental Figure 1

**Gene Curation Approach for CVD Gene Panel.** The evidence evaluated was dependent on the classification of the GDR and whether the GDR source was from ClinGen or a group that uses the ClinGen Gene-Disease Validity framework for gene curation. GDRs were either verified or curated using an expedited process with select GDRs subject to a preliminary literature search to assess the need to update the available curation. GDRs that met the threshold of a strong or definitive GDR were included in the CVD gene panel.

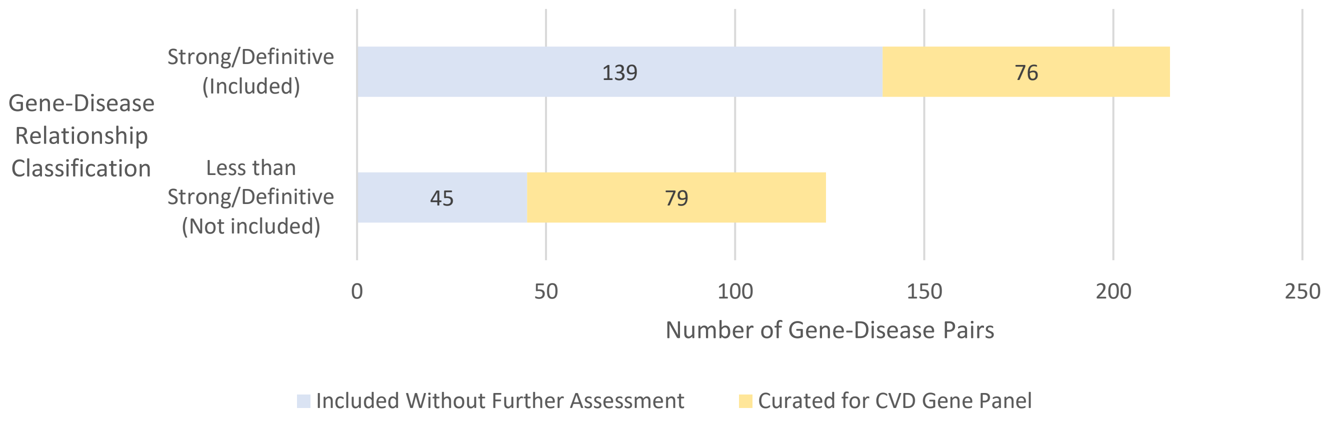

#### Supplemental Figure 2.

**Outcomes of Gene-Disease Curation by Curation Source.** A threshold of a strong or definitive GDR classification was set as the threshold for inclusion in the CVD gene panel. Based on the classification of the GDR and the GDR source, gene-disease pairs were either included or not included without further assessment or curated using a modified version of the ClinGen Gene-Disease Validity Framework.

#### Polygenic Risk Score for Coronary Artery Disease

**Elevated**

This individual has an elevated risk to develop **coronary artery disease (CAD)** based on their calculated polygenic risk score (PRS).

This individual belongs to a group with similar genetic ancestry that has a 2-fold increased risk to develop CAD compared to the remainder of the individuals in that ancestry group. This group's calculated risk to develop CAD based on their PRS is between **1.7-fold** and **3.3-fold** increased.

The methods to calculate PRS are continuing to improve, and some uncertainty remains in terms of how well these scores predict the risk of developing disease. This risk is an estimate of this individual's chance to develop CAD, but the actual risk of disease may be greater or smaller. For example, the overall risk of developing CAD is also affected by genetic variants which may not be included in the PRS calculation, as well as medical history, family history, and lifestyle factors not accounted for by this test. Clinical correlation is recommended.

#### Methods and Limitations

A PRS for CAD was calculated based on a set of at least 123,622 genetic variants evaluated in the submitted specimen. Ancestry-specific CAD PRSs developed by Allelica, Inc. were used in this test to calculate this individual's risk for CAD (PMID: 33677976; PMID: 35513724; Busby GB, et al., 2022 retrieved from https://saas.allelica.com/allelica_manuscript_CAD_multiancestry.pdf). Ancestry is calculated using iAdmix (PMID: 25592880). Computed ancestry groups include African, American Admixed, East Asian, European, and South Asian. Individuals are assigned to a single genetic ancestry group if they have >=80% ancestry from one of these five groups. PRSs for admixed individuals are calculated using the weighted average of PRSs from each of their underlying ancestry groups as listed above.

Ancestry-specific PRSs are based on the combination of one or more ancestry-specific Genome Wide Association Studies (GWAS). The ancestry specific PRSs used in this test were the highest performing of those evaluated in each ancestry group based on odds ratio per standard deviation (ORxSTD)). Each PRS is calculated based on a defined set of single nucleotide polymorphisms (SNPs). The sum of the risk alleles for each SNP is weighted by the corresponding effect size from the PRS panel.

Relative risk of CAD was calculated by identifying the PRS percentile above which there is a 2-fold risk compared to the remainder of the population in that ancestry group [white paper]. Elevated risk for CAD on this test is defined as anyone with a PRS above the 2-fold threshold.

The results of this PRS test, performed in a high complexity Clinical Laboratory Improvement Amendment (CLIA) laboratory under 42 CFR 493(b)(2) exception, are intended for research purposes only and are not validated for the diagnosis, prevention, or treatment of a specific disease. The risk estimate provided is intended to represent an individual's risk using common genetic variants alone. All risk estimation is approximate and based on previously analyzed cohorts. Being identified as at elevated risk is not a diagnosis and does not necessarily indicate that the individual will develop CAD.

#### Supplemental Figure 3.

**Example PRS report content for an individual with an elevated risk for CAD based on their PRS.** The CAD PRS calculates the tested individual's genetic ancestry and applies the corresponding ancestry specific PRS. Elevated risk of CAD is defined as a 2-fold increased relative risk of developing CAD compared to the remainder of individuals in that ancestry group. The CAD PRS report highlights uncertainty about the ability of PRS to predict disease, consideration of genetic variants which may not be included in the PRS calculation, and the importance of medical history, family history, and lifestyle factors.
